# Health-Related Quality of Life and Coping Strategies adopted by COVID-19 survivors: A nationwide cross-sectional study in Bangladesh

**DOI:** 10.1101/2022.04.30.22274514

**Authors:** Mohammad Anwar Hossain, Rubayet Shafin, Md. Shahoriar Ahmed, Md. Shohag Rana, Lori Maria Walton, Veena Raigangar, Tasnim Ara, Md. Aminul Haque Rasel, Mohammad Sohrab Hossain, Md. Feroz Kabir, Mir Raihanul Islam, Md. Nazmul Hasan, Delowar Hossain, Farjana Sharmin Rumana, Iqbal Kabir Jahid

**Affiliations:** Department of Microbiology, Jashore University of Science & Technology (JUST), Jashore, Bangladesh; Department of Physiotherapy, Centre for the Rehabilitation of the Paralysed (CRP), Savar, Dhaka, Bangladesh; Department of Physiotherapy, Bangladesh Health professions Institute (BHPI), Dhaka, Bangladesh; Department of Physical Therapy, School of Health Sciences, University of Scranton, Pennsylvania, USA; Department of Physiotherapy, College of Health Sciences, University of Sharjah, Sharjah, U.A.E; Institute of Statistical Research and Training, University of Dhaka; Department of Physiotherapy & Rehabilitation, Jashore University of Science & Technology (JUST), Jashore, Bangladesh; Poverty, Health and Nutrition Division, International Food Policy Research Institute; Department of Physiotherapy & Rehabilitation, Enam Medical College Hospital

**Keywords:** QoL, Coping strategies, COVID-19, Bangladesh

## Abstract

This study aims to investigate the health-related quality of life and coping strategies among COVID-19 survivors in Bangladesh.

**Methods:** This is a cross-sectional study of 2198 adult, COVID-19 survivors living in Bangladesh. Data were collected from previously diagnosed COVID-19 participants (confirmed by an RT-PCR test) via door-to-door interviews in the eight different divisions in Bangladesh. For data collection, Bengali translated Brief COPE inventory and WHO Brief Quality of Life (WHO-QOLBREF) questionnaires were used. The data collection period was from June 2020 to March 2021.

**Results:** Males 72.38% (1591) were more affected by COVID-19 than females 27.62% (607). Age showed significant correlations (p<0.005) with physical, psychological and social relationships; whereas, gender showed only significant correlation with physical health (p<0.001). Marital status, occupation, living area, and co-morbidities showed significant co-relation with all four domains of QoL (p<0.001). Education and affected family members showed significant correlation with physical and social relationship (p<0.001). However, smoking habit showed significant correlations with both social relationship and environment (p<0.001). Age and marital status showed a significant correlation with avoidant coping strategy (p<0.001); whereas gender and co-morbidities showed significant correlation with problem focused coping strategies (p<0.001). Educational qualification, occupation and living area showed significant correlation with all three coping strategies (p<0.001).

**Conclusion:** Survivors of COVID-19 showed mixed types of coping strategies; however, the predominant coping strategy was avoidant coping, followed by problem focused coping, with emotion focused coping reported as the least prevalent. Marital status, occupation, living area and co-morbidities showed a greater effect on QoL in all participants. This study represents the real scenario of nationwide health associated quality of life and coping strategy during and beyond the Delta pandemic.

## Background

In Bangladesh, the COVID-19 pandemic has progressed rapidly overtime and the burden of the Delta variant entering from neighboring countries [1], in addition to lack of resources within Bangladesh, low vaccine availability, affordability, accessibility and implementation have added to the country’s devastating COVID-19 infection rates and death rates. As the country prepared for its fourth wave, the infection rate was estimated to be over 30% on July 26, 2021[2]. As of October 29, 2021, in Bangladesh, the total samples tested were 10,301,593 of which 1,568,857 confirmed cases and 27,847 deaths [3]. The increased death in Bangladesh during this period was attributed due to the second wave, initially by B.1351 [4] and B1.617.2 [5]. With 150 nations, since March 18, 2021, Bangladesh suspended all academic institutions [6] and from March 26, 2021, the Bangladeshi Government encouraged people to stay home to prevent the rapid spread of COVID-19. This long-time, infrequent lockdown that started from March 10, 2020 [7], coupled with the severity of COVID-19 and its impact on individual’s social and mental health, with a high impact on everyday stress and anxiety levels for the general population. All these factors had a substantial negative impact on the Quality of life (QoL) [8-10].

Quality of life is a broad term and represents one’s overall physical, mental, social, and environmental satisfaction. Due to the loss of lives and livelihoods, COVID-19 has exacerbated psychosocial and socioeconomic insecurity among poor people by causing price hike of basic products, restriction of informal education, and the risk of a serious socio-economic and health crisis [11]. However, due to the shutdown of exports and imports, many people lost their jobs (for example, garment workers, corporate office employees, and foreign revenue declines) further affecting the quality of life (QoL) for people already struggling economically prior to the onset of COVID-19 [12]. Humans have shown great capacity for developing a variety of coping mechanisms for survival during and after catastrophic events. However, the extra burden of poverty on people during a catastrophic event has been shown to have cumulative negative impacts on the psychological coping strategies for people over long periods of time [13]. Coping methods are emotion-driven efforts to handle stress that has been linked to improved mental health and are necessary components to healing from trauma [14]. Studies have shown that the coping method adopted by individuals has a significant impact on how they experience anxiety and process behavioral responses, accordingly [15]. Communication, avoidance and activities are some of the methods being used as Coping strategies. From the definition COPING is “Efforts to prevent or diminish threat, harm, and loss, or to reduce the distress that is often associated with those experiences” [16] which can be described as the broad terms “Approach-an issue is solved by controlling stress” and “Avoidant-a problem is solved by avoiding stress by reducing unpleasant emotions”. Scholarly evidence shows that Approach Coping Strategy (APC) is more common in the Bangladeshi community than Avoidance Coping Strategy (AVC) [17]. To the best of our knowledge, there is a scarcity of empirical evidence concerning the effects of COVID-19 on coping and QoL among the patients recovering from this infectious disease. Therefore, the purpose of this research was compared between behavioral aspects of COPING strategies and QoL among COVID-19 affected populations in Bangladesh.

## Methodology

### Study design

This was a cross-sectional study of 2198 adult COVID-19 survivors collected from 14392 COVID-19 positive cases across all divisions of Bangladesh from the time frame between October, 2020 to March, 2021. All the participants tested positive or negative through RT-PCR nasopharyngeal swab under the national surveillance systems of COVID-19 located at the Directorate of General of Health Services (DGHS) in various laboratories throughout Bangladesh [18]. The RT-PCR test for SARS-CoV-2 has been documented as the gold standard and most of the countries, including Bangladesh, are using the RT-PCR for diagnosis the COVID-19 [19]. The inclusion factors for this study were age 18 years and above, participants with persistent secondary complications after a positive diagnosis, and those who reported difficulties undertaking usual daily activities. Exclusion criteria were persistent fever, too sick to participate, mental instability, decline consent, and those who were untraceable.

### Sample size

The sample size calculation was performed using “EPI INFO” software version 7.4.2.0 developed by the Center for Disease Control in the US. For the calculation. The reference figure of 1,562,359 was used (i.e., The total number of COVID-19 positive cases reported up to October 2021) [2] with a cluster figure of eight (the number of administrative divisions in Bangladesh) A calculation was then made with 50% of expected frequency, 5% margin of error, and 1.0 design effect. The sample size was generated as a minimum of 1520 with a minimum of 168 samples per division.

### Study procedure

Data were collected through appointed trained assessors from the Centre for the Rehabilitation of the Paralyzed (CRP). The research team initially reviewed the materials from WHO and made a framework of questionnaire and drafted individual questions through an interactive process of zoom meeting which is under review in other journals by the help of different disciplines such as Microbiologist, Physicians, physiotherapist. The questionnaire were initially drafted in English but later translated by researchers who had good knowledge in both languages. Before data collection all assessors were comprehensively trained by the principal author regarding study protocols, precaution, adverse events, aims, ethical considerations, questionnaires and the possible outcomes. A pilot study was conducted with 20 participants, with face-to-face data collection and was undertaken at a convenient scheduled time for participants, after taking written consent from the participants. Informed consent and questionnaires were provided in paper format, with consent read aloud for every participant in their native language to assure full comprehension. During data collection, all the assessors adhered to the COVID-19 preventive precautions by utilizing personal protective equipment (PPE) and general health regulations set forth by the Bangladesh Government. Data were collected in paper format and then transferred into Excel Workbook for external data audit. After completion of the data audit, the data was analyzed in SPSS, version 20.0.

### Data collection and Questionnaire

A phone call follow-up was conducted with all participants (N=13,222) for any secondary complications after receiving a negative test result for COVID-19. A total 2310 participants with secondary complications provided consent and completed the questionnaire. The first part of the questionnaire provided socio-demographic information, the second part provided comorbidity information, blood group and rhesus status, date of COVID-19 positive test, date of COVID-19 negative test, presenting symptoms during COVID-19 illness, persisting COVID-19 symptoms, and treatment received during COVID-19 illness and the third part provided the cardio-respiratory parameter measurements and included: resting heart rate (HR); blood oxygen saturation levels (Spo2); systolic and diastolic blood pressure; inspiratory and expiratory lung volumes; and maximal oxygen consumption (Vo2max). All surveys were translated from English to Bengali language and back translated from Bengali to English language, and the language validation process was followed as per WHO guidance [20]. The Brief-COPE is a frequently used self-reported questionnaire that was developed to assess a broad range of coping strategies. It has 28 items questionnaires that describes the COPING responses in three domains (problem, emotion and avoidant focused). Each item in each domain is scored from possible options on an ordinal scale from one to four. The World Health Organization Quality of Life-BREF scale was used to determine QOL. The WHOQOL-BREF is a 26-item scale that is used to assess people’s quality of life. It is an abbreviated version of the WHOQOL-100 scale. It consists of four domains as well as a general health domain. Physical health (7 items), psychological health (6 items), social relationships (3 items), and the environment (8 domains). The final two items are from the general health domain, which enables respondents to score their overall satisfaction with their health and quality of life. The scale items are graded on a five-point Likert scale ranging from 1 (very dissatisfied/very poor) to 5 (very satisfied/very good), with higher scores indicating better quality of life [21]. Large values of KMO statistic (>0.8) for both WHO-QoL and Brief Cope questionnaire indicated that the sample was suitable for factor analysis. On the other hand, the reliability was determined by calculating Cronbach’s α coefficient. The coefficient was measured as 0.716 and 0.886 respectively, well above the minimum accepted threshold of 0.70 [22].

### Statistical Testing

Data were analyzed using the Statistical Package for Social Science (SPSS) version 20.0 [23]. The Kaiser-Meyer-Olkin (KMO) analysis were done between WHO QoL and Coping for data adequacy and normality for factor analysis and descriptive analysis performed for parametric socio-demographic, dependent variable and health and co-morbidities of the respondents (Table 1). In addition, multivariate analysis of variance (One-way MANOVA) statistics were performed for dependent variables between QoL and coping strategies (Table 2). Population distribution is shown in the Box plot (Figure: 2). The alpha value was set as p<0.05.

**Table 1:**
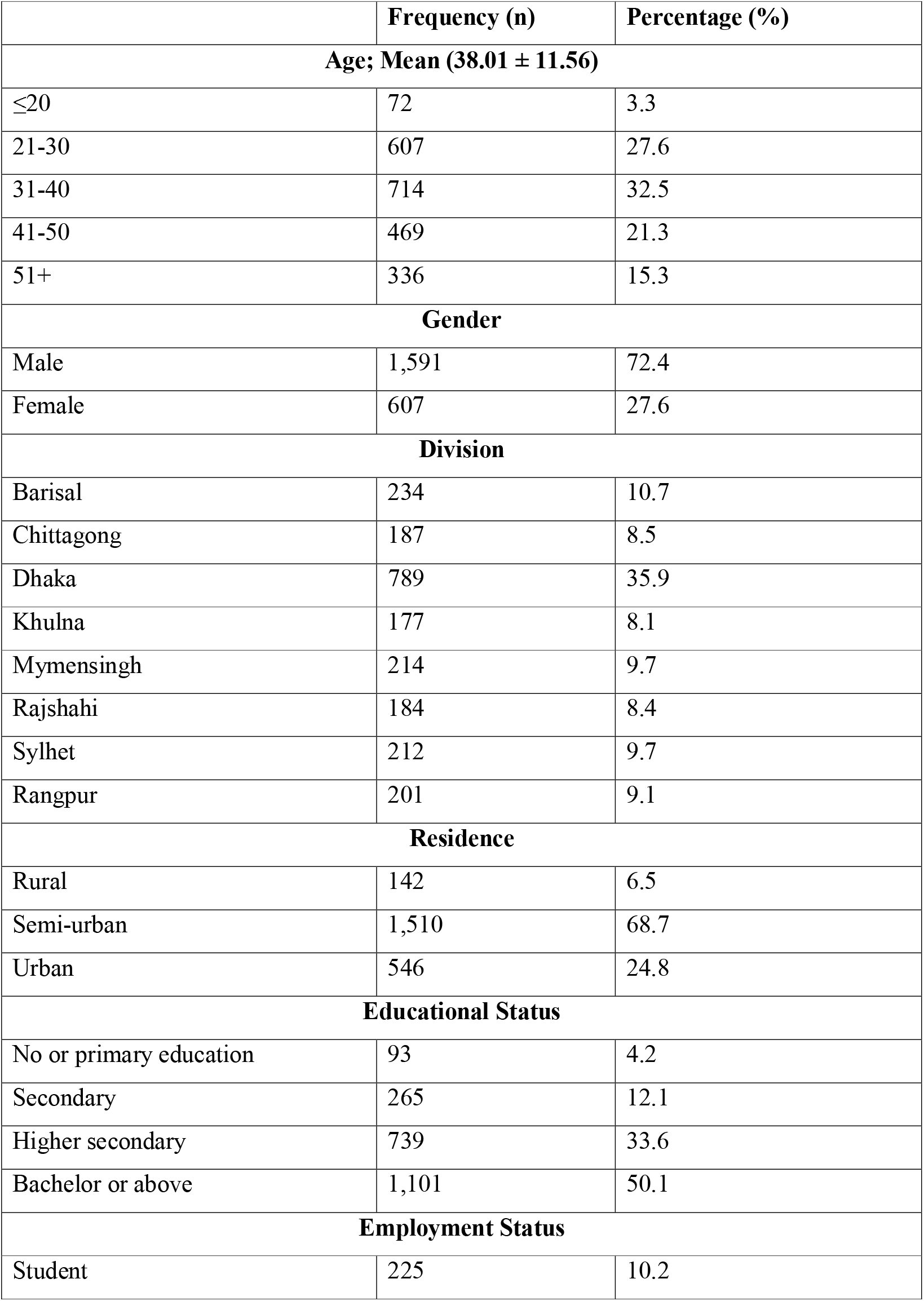
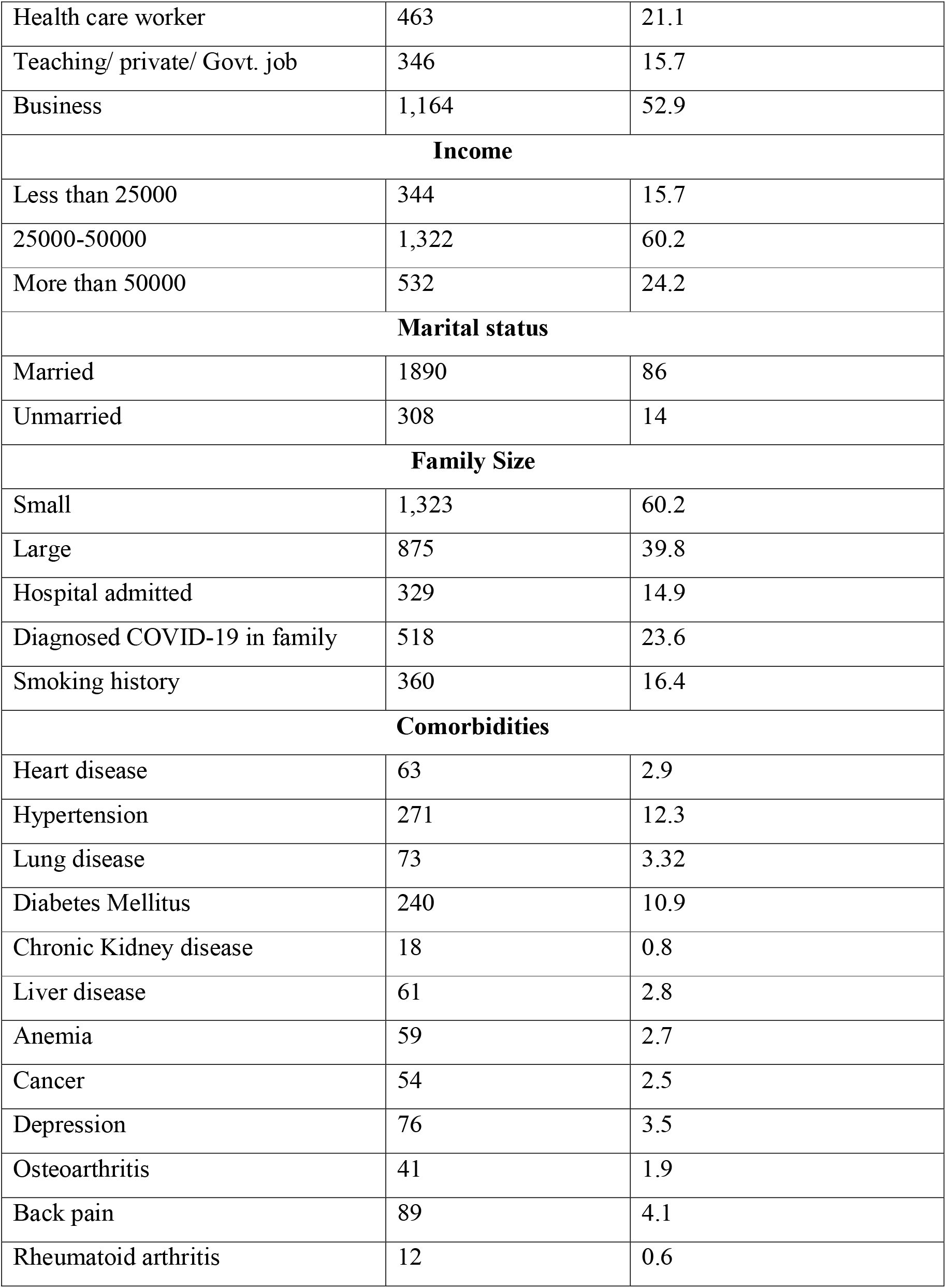
Demographic, health situation and comorbidities characteristics of the analytic sample.

|  | Frequency (n) | Percentage (%) |
| --- | --- | --- |
| **Age; Mean (38.01 ± 11.56)** |  |  |
| ≤20 | 72 | 3.3 |
| 21-30 | 607 | 27.6 |
| 31-40 | 714 | 32.5 |
| 41-50 | 469 | 21.3 |
| 51+ | 336 | 15.3 |
| **Gender** |  |  |
| Male | 1,591 | 72.4 |
| Female | 607 | 27.6 |
| **Division** |  |  |
| Barisal | 234 | 10.7 |
| Chittagong | 187 | 8.5 |
| Dhaka | 789 | 35.9 |
| Khulna | 177 | 8.1 |
| Mymensingh | 214 | 9.7 |
| Rajshahi | 184 | 8.4 |
| Sylhet | 212 | 9.7 |
| Rangpur | 201 | 9.1 |
| **Residence** |  |  |
| Rural | 142 | 6.5 |
| Semi-urban | 1,510 | 68.7 |
| Urban | 546 | 24.8 |
| **Educational Status** |  |  |
| No or primary education | 93 | 4.2 |
| Secondary | 265 | 12.1 |
| Higher secondary | 739 | 33.6 |
| Bachelor or above | 1,101 | 50.1 |
| **Employment Status** |  |  |
| Student | 225 | 10.2 |
| Health care worker | 463 | 21.1 |
| Teaching/ private/ Govt. job | 346 | 15.7 |
| Business | 1,164 | 52.9 |
| **Income** |  |  |
| Less than 25000 | 344 | 15.7 |
| 25000-50000 | 1,322 | 60.2 |
| More than 50000 | 532 | 24.2 |
| **Marital status** |  |  |
| Married | 1890 | 86 |
| Unmarried | 308 | 14 |
| **Family Size** |  |  |
| Small | 1,323 | 60.2 |
| Large | 875 | 39.8 |
| Hospital admitted | 329 | 14.9 |
| Diagnosed COVID-19 in family | 518 | 23.6 |
| Smoking history | 360 | 16.4 |
| **Comorbidities** |  |  |
| Heart disease | 63 | 2.9 |
| Hypertension | 271 | 12.3 |
| Lung disease | 73 | 3.32 |
| Diabetes Mellitus | 240 | 10.9 |
| Chronic Kidney disease | 18 | 0.8 |
| Liver disease | 61 | 2.8 |
| Anemia | 59 | 2.7 |
| Cancer | 54 | 2.5 |
| Depression | 76 | 3.5 |
| Osteoarthritis | 41 | 1.9 |
| Back pain | 89 | 4.1 |
| Rheumatoid arthritis | 12 | 0.6 |

**Table 2:** One-way MANOVA in between demographic variables with WHO quality of life

| Variables | Physical health |  |  | Psychological |  |  | Social relationship |  |  | Environmental |  |  |
| --- | --- | --- | --- | --- | --- | --- | --- | --- | --- | --- | --- | --- |
|  | Mean±SE | F (p) | Partial<br>η<sup>2</sup> | Mean±SE | F (p) | Partial<br>η<sup>2</sup> | Mean±SE | F (p) | Partial<br>η<sup>2</sup> | Mean±SE | F (p) | Partial<br>η<sup>2</sup> |
| Age |  |  |  |  |  |  |  |  |  |  |  |  |
| less than<br>equal 20 | 93.5±1.1 | 22.9*** | .04 | 85.9±1.1 | 43.4*** | .073 | 41.1±.5 | 26.6*** | .046 | 94.8±.8 | .9 | .002 |
| 21-30 years | 93.6±.4 |  |  | 80.2±.4 |  |  | 40±.2 |  |  | 95.5±.3 |  |  |
| 31-40 years | 94±.3 |  |  | 78.5±.3 |  |  | 39.5±.2 |  |  | 95.5±.2 |  |  |
| 41-50 years | 91.9±.4 |  |  | 76.2±.4 |  |  | 38.2±.2 |  |  | 95.1±.3 |  |  |
| more than<br>equal 51 | 88.8±.5 |  |  | 73.8±.5 |  |  | 37.4±.2 |  |  | 94.9±.4 |  |  |
| less than<br>equal 20 | 93.5±1.1 |  |  | 85.9±1.1 |  |  | 41.1±.5 |  |  | 94.8±.8 |  |  |
| Gender |  |  |  |  |  |  |  |  |  |  |  |  |
| Female | 90.6±.4 | 41.1*** | .018 | 77.1±.4 | 7.1 | .003 | 38.9±.2 | 1.6 | .001 | 95±.3 | 1.5 | .001 |
| Male | 93.4±.2 |  |  | 78.4±.2 |  |  | 39.2±.1 |  |  | 95.4±.2 |  |  |
| Marital status |  |  |  |  |  |  |  |  |  |  |  |  |
| Married | 92.4±.2 | 7.1** | .003 | 77.1±.2 | 131.1*** | .056 | 38.8±.1 | 55.6*** | .025 | 95.3±.2 | .05 | .000 |
| Unmarried | 93.9±.5 |  |  | 83.7±.5 |  |  | 40.9±.3 |  |  | 95.2±.4 |  |  |
| Education |  |  |  |  |  |  |  |  |  |  |  |  |
| No formal Education | 86.4±1.4 | 11.8*** | .021 | 76.9±1.6 | 6.1*** | .011 | 40.3±.8 | 14.7*** | .026 | 90.3±1.1 | 19.2*** | .034 |
| Primary Education | 93.3±1.2 |  |  | 76.8±1.2 |  |  | 37.5±.6 |  |  | 93.3±.9 |  |  |
| Secondary Education | 91.7±.6 |  |  | 78.4±.6 |  |  | 38.8±.3 |  |  | 93.3±.4 |  |  |
| Higher secondary Education | 91.5±.3 |  |  | 76.7±.4 |  |  | 38.3±.2 |  |  | 95.1±.2 |  |  |
| Bachelor or above | 93.7±.3 |  |  | 78.9±.3 |  |  | 39.8±.1 |  |  | 96.2±.2 |  |  |
| Occupation |  |  |  |  |  |  |  |  |  |  |  |  |
| Students | 94.4±.6 | 13.1*** | .040 | 85.4±.6 | 34.6*** | .100 | 41.1±.3 | 18.3*** | .056 | 95.7±.4 | 6.9*** | .022 |
| Health care professionals | 87.5±.8 |  |  | 74.3±.8 |  |  | 40.4±.4 |  |  | 96.1±.6 |  |  |
| Law enforcement agency | 93.1±.9 |  |  | 82.2±.9 |  |  | 41.6±.5 |  |  | 94.2±.7 |  |  |
| Housewife | 90.1±.6 |  |  | 76.4±.6 |  |  | 38.6±.3 |  |  | 94.2±.4 |  |  |
| Government jobs | 91.6±.5 |  |  | 75.8±.6 |  |  | 39.2±.3 |  |  | 94.9±.4 |  |  |
| Private jobs | 93.6±.3 |  |  | 77.9±.3 |  |  | 38.7±.1 |  |  | 95.8±.2 |  |  |
| Farmer and worker | 91.4±1.2 |  |  | 72.2±1.3 |  |  | 36.3±.7 |  |  | 90.3±.9 |  |  |
| Unemployed | 91.2±1.5 |  |  | 72.9±1.5 |  |  | 36.5±.8 |  |  | 93.9±1.1 |  |  |
| Living area |  |  |  |  |  |  |  |  |  |  |  |  |
| Rural | 94.4±.8 | 8.8*** | .008 | 79.9±.8 | 35.3*** | .031 | 40.5±.4 | 114.2*** | .094 | 90.5±.5 | 47.1*** | .041 |
| Semi Urban | 92.9±.2 |  |  | 76.9±.2 |  |  | 38.2±.1 |  |  | 95.9±.2 |  |  |
| Urban | 91.3±.4 |  |  | 80.7±.4 |  |  | 41.4±.2 |  |  | 94.9±.3 |  |  |
| Family members affected |  |  |  |  |  |  |  |  |  |  |  |  |
| Affected family members | 91.1±.4 | 19.4*** | .009 | 78.2±.4 | .2 | .000 | 40.4±.2 | 48.5*** | .022 | 94.5±.3 | 10.0 | .005 |
| Unaffected family members | 93.1±.2 |  |  | 77.9±.2 |  |  | 38.7±.1 |  |  | 95.6±.2 |  |  |
| Smoking history |  |  |  |  |  |  |  |  |  |  |  |  |
| Smokers | 92.5±.5 | .1 | .000 | 78.2±.5 | .1 | .000 | 40.2±.2 | 22.8*** | .010 | 93.1±.3 | 48.6*** | .022 |
| Non smoker | 92.6±.2 |  |  | 77.9±.2 |  |  | 38.9±.1 |  |  | 95.7±.2 |  |  |
| Comorbidities |  |  |  |  |  |  |  |  |  |  |  |  |
| No comorbidity | 93.9±.2 | 60.4*** | .076 | 79.1±.2 | 43.7*** | .056 | 39.2±.1 | 6.7*** | .009 | 95.6±.2 | 11.2*** | .015 |
| one<br>comorbidity | 90.8±.5 |  |  | 77.4±.6 |  |  | 39.8±.3 |  |  | 95.6±.4 |  |  |
| two<br>comorbidities | 86.9±.7 |  |  | 72.1±.7 |  |  | 37.9±.4 |  |  | 94.3±.5 |  |  |
| more than<br>two<br>comorbidities | 86.7±.8 |  |  | 72.8±.8 |  |  | 38.4±.4 |  |  | 92.3±.6 |  |  |

Significant relationship values with a minimum of 5% margin of error are bolded and marked as \* p<.05, \*\* p<.01, \*\*\* p<.001

## Results

### Demographic characteristics of the analytic sample

The socio-demographic characteristics of the analytical sample are demonstrated in Table 1. A total 2198 participants aged 18 years to 86 years of age (38±11.6 years) responded to the survey. Most of the respondents 32.5% (n=714) were from the age group (31–40 years). Male respondents were 72.4% (n=1591) and female 27.6% (n=607). Regional disaggregation of the samples showed that most of the respondents were 35.9% (n=789) from the Dhaka division and over two third of them were 68.7% (n=1510) living in the semi-urban areas. More than half of the participants were (52.9%, n=1164) involved in business activities, and 50.1% (n=1101) reported completion of a Bachelor’s Degree. Around 15.7% (n=1322) of the respondents reported income less than 25000TK per month and the majority (84.9%, n=1868) of the respondents reported being married. Almost 60.2% (n=1323) of the sample belong to a small family size. A small number of the participants, around 16.4% (n=360) reported were previous smoking status (Table 1).

### Comorbidities of the respondents

Approximately 15% (n=329) of the respondents were admitted into hospitals and most of them, 85% (n=1869) were not hospitalized. Almost one-fourth of the respondents have at least one family member who was diagnosed with COVID-19 23.6% (n=518). Comorbidities represented included: hypertension was found to have the highest prevalence at 12.33% (n=271) followed by Diabetes Mellitus reported as second highest at 10.9% (n=240). All other comorbidities reported almost the same prevalence rate, including: heart disease at 2.9% (n=63), lung disease at 3.3% (n=73), chronic kidney disease at 0.8% (n=18), liver disease at 2.8% (n=61), anemia or other blood diseases at 2.7% (n=59), cancer at 2.5% (n=54), depression at 3.5% (n=76), osteoarthritis at 1.9% (n=41), back pain at 4.1% (n=89), and rheumatoid arthritis 0.6% (n=12) (Table 1).

The differences in the usage of the three coping strategies were tested using one-way Multivariate analysis of variance (MANOVA) procedure. The differences in the usage of the three coping strategies were tested using one-way Multivariate analysis of variance (MANOVA) procedure.

### Relation of demographic with quality of life

There was a significant correlation of sociodemographic and quality of life of COVID-19 survivors (Table 2). Age and gender showed significant correlation with physical (p<0.001), psychological (p<0.001, 0.007), and social (p<0.001) but did not significantly correlate with environment. Division, residence, education and employment showed significant correlation with all four domains (p<0.001). Income showed positive correlation with physical (p<0.001), psychological (p<0.001) and environmental (p<0.001) but not significantly correlate with social domain. Marital status shows significant correlation with psychological (p<0.001) and social (p<0.001) but no significant correlation found with physical and environmental domain. Family size shows significant correlation with physical and psychological (p<0.001) but not with social and environmental. Hospitalized due COVID-19 and comorbidity shows positive correlation with all four domains (p<0.001). Diagnosed COVID-19 in the family shows only significant association with physical (p<0.001) domain. Smoker shows positive co-relation with social (p<0.001) and non-smoker showed positive association with environment (p<0.001) (Table 2).

### Relation of demographic with COPING strategies

From Table 3 it is apparent that age had a significant correlation with Avoidant coping strategy (p<.05). From the age category, respondents who was less than 20 years old had higher mean score (6.69±3.66) to the avoidant coping. Gender had significant correlation with problem focused coping strategy (p<.005) where male participants had higher mean scores than female (7.52±5.88) to problem focused coping. Marital status had significant relation with emotion focused (p<.05) and avoidant coping (p<.001) strategy and among the marital status category unmarried participants had highest mean score to the emotion focused (13.08±.17) and avoidant coping (6.52±.08) strategies respectively. Educational qualification had significant correlation with all three coping strategies (p<.001p<.001p<.001). From the respondents, who were bachelor or above had highest mean score to the problem focused coping (7.98±.591) whereas respondents who completed higher secondary had highest mean score to the emotion focused coping and avoidant coping (12.81±.715; 6.78±.328) strategies.

**Table 3:**
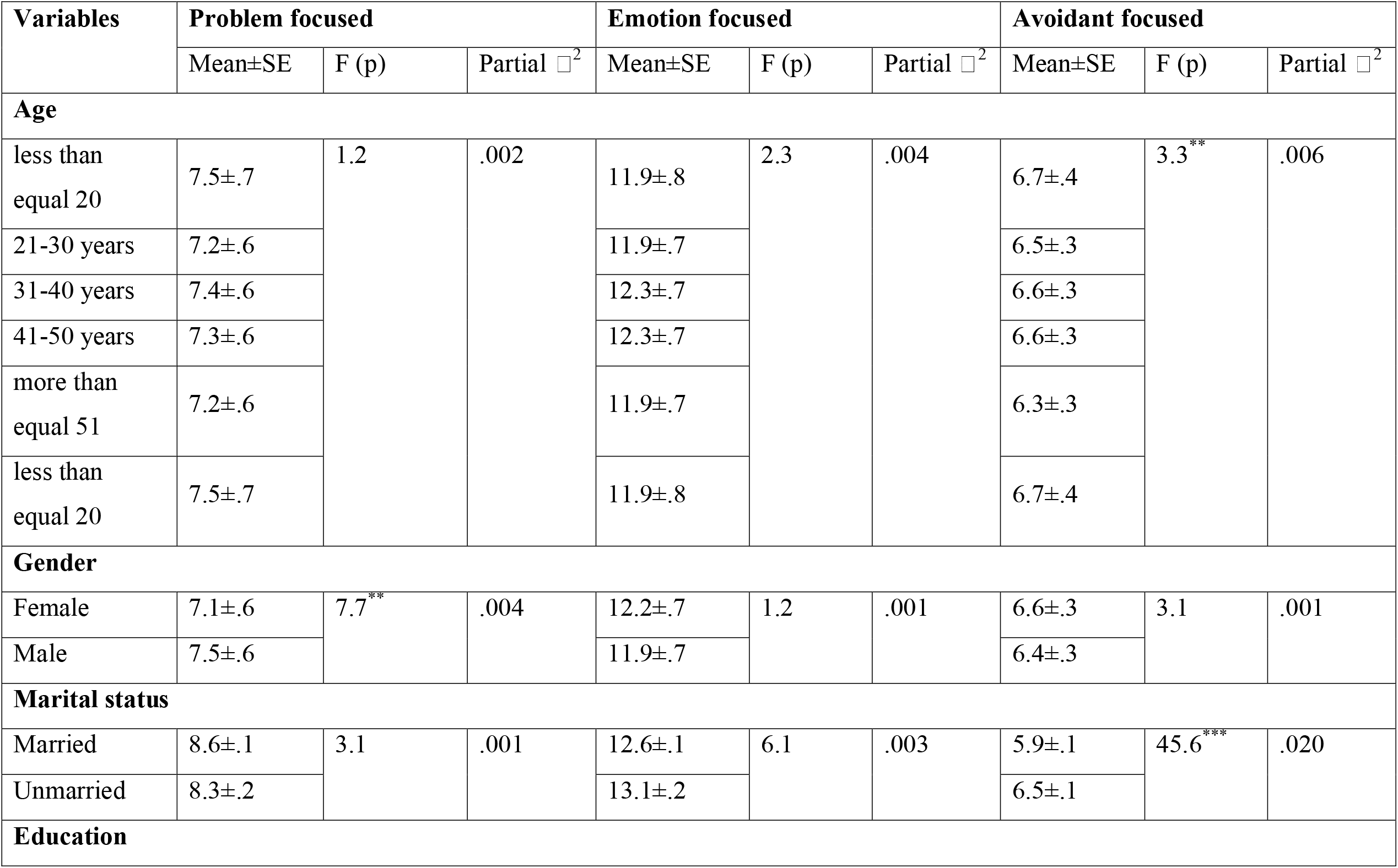
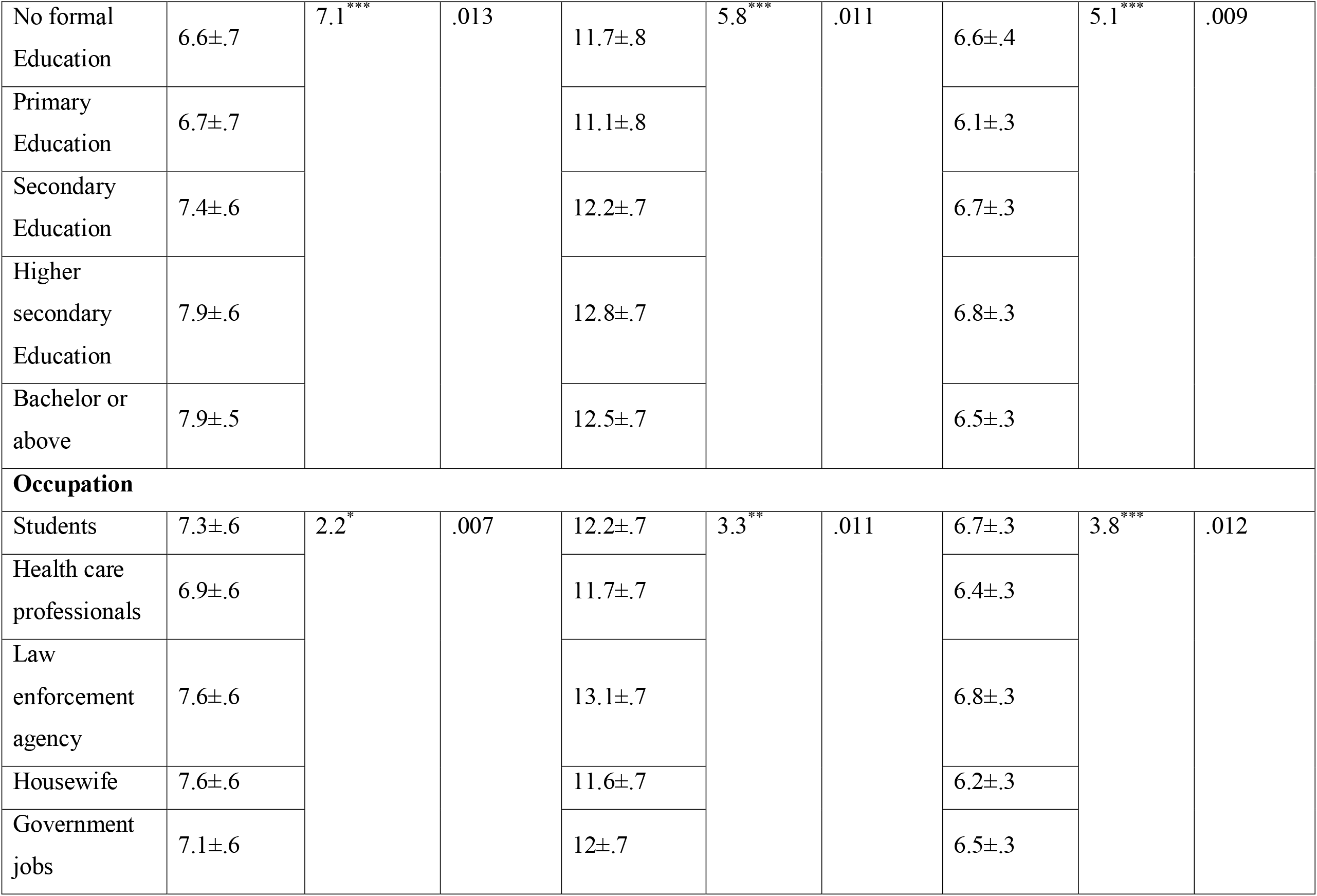
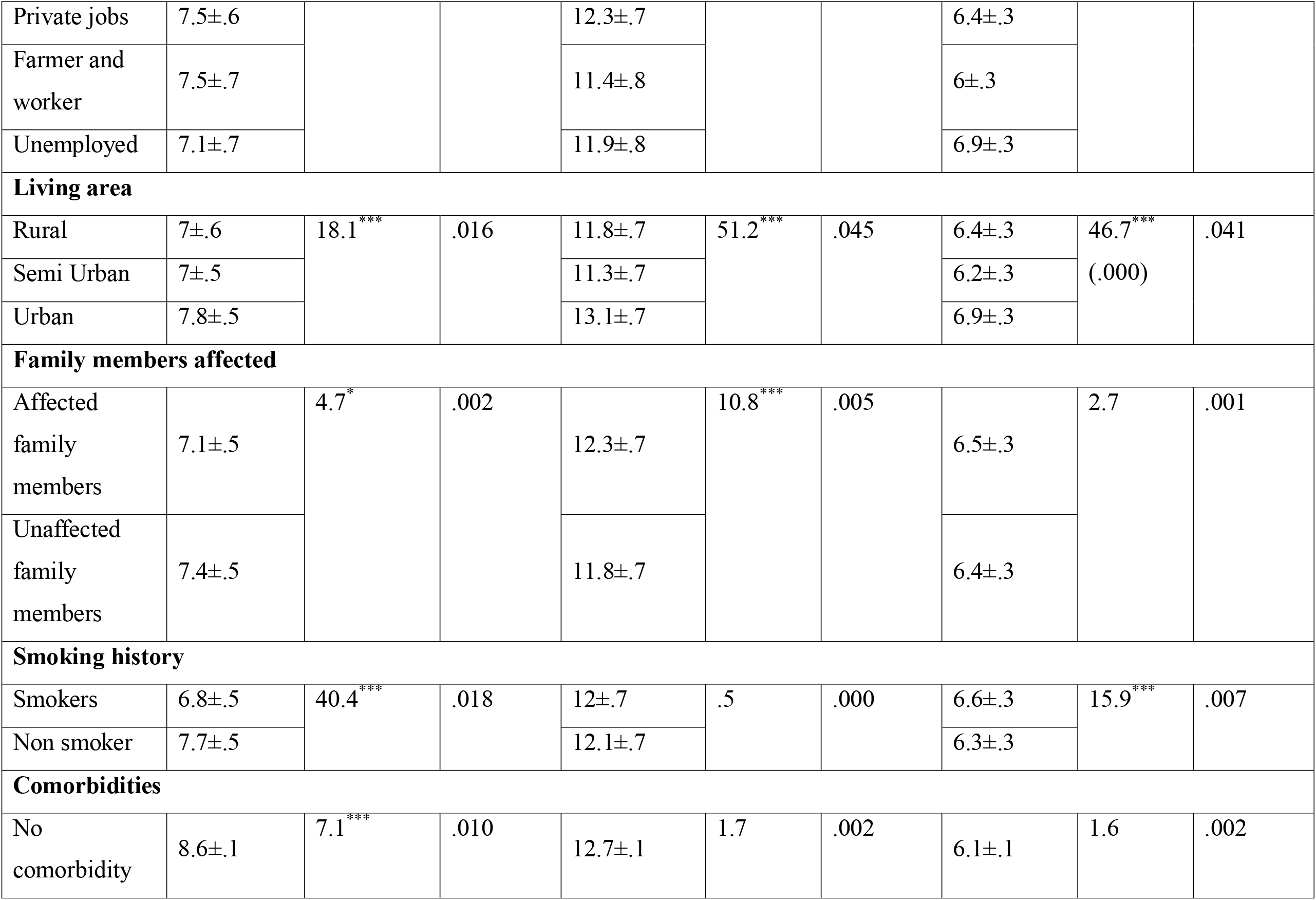
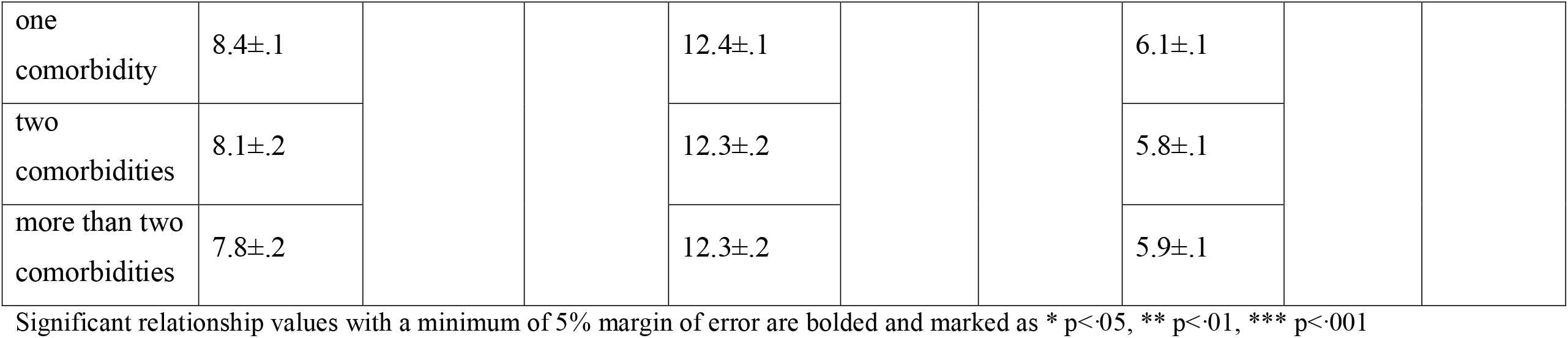
One-way MANOVA in between demographic variables with COPING strategies

Occupational status had also significant correlation with all three coping strategies (p<.05; p<.001 p<.001). From the occupational categories housewives had highest mean score (7.60±.612) to problem focused coping strategy adoption. Respondents who were from law-enforcement agency had highest mean score (13.17±.768) to the emotion focused coping and those who were unemployed had highest mean score to the avoidant coping strategy (6.96±.389). Living area shows significant correlation with all three coping strategies (p<.001; p<.001; p<.001) (Table: 3).

Some weak to moderate correlations were found between COPING and the QoL domains. Also, coping factors correlated with several QoL domains: Problem focused coping was associated with psychological (r = .165, p <0.001 and social relation (r = 0.061, p > 0.001). Emotion focused coping correlated with psychological (r = .104, p <0.001) and social relation (r = .150, p > 0.001) and negatively associated with physical health (r=-.090, p <0.001) and environment (r = -.236, p > 0.001). Avoidant coping was positively associated with psychological (r = 0.56, p > 0.001) but negatively associated with physical health (r=-.220, p <0.001) and environment construct? (r = -.217, p > 0.001) (Table 4).

**Table 4:** Correlation between COPING and QoL by Pearson correlation.

|  |  | Problem<br>focused | Emotion<br>focused | Avoidant<br>focused | Physical<br>health | Psychological | Social<br>relation | Environment |
| --- | --- | --- | --- | --- | --- | --- | --- | --- |
| Problem<br>focused | Pearson<br>Correlation | 1 | .678 | .330 | .017 | .165 | .061 | -.027 |
|  | Sig. |  | .000 | .000 | .422 | .000*** | .004** | .198 |
| Emotion<br>focused | Pearson<br>Correlation |  | 1 | .572 | -.090 | .104 | .150 | -.236 |
|  | Sig. |  |  | .000 | .000 | .000*** | .000*** | .000 |
| Avoidant<br>focused | Pearson<br>Correlation |  |  | 1 | -.220 | .056 | .039 | -.217 |
|  | Sig. |  |  |  | .000 | .009** | .065 | .000 |
| Physical<br>health | Pearson<br>Correlation |  |  |  | 1 | .559 | .239 | .315 |
|  | Sig. |  |  |  |  | .000 | .000 | .000 |
| Psychologi<br>cal | Pearson<br>Correlation |  |  |  |  | 1 | .243 | .289 |
|  | Sig. |  |  |  |  |  | .000 | .000 |
| Social<br>relation | Pearson<br>Correlation |  |  |  |  |  | 1 | .058 |
|  | Sig. |  |  |  |  |  |  | .006 |
| Environme<br>nt | Pearson<br>Correlation |  |  |  |  |  |  | 1 |
|  | Sig. |  |  |  |  |  |  |  |

Significant relationship values with a minimum of 5% margin of error are bolded and marked as \*
$p<.05$ , \*\* $p<.01$ , \*\*\* $p<.001$

From the boxplot Fig. 2 (A) In quality-of-life domains, participants less than 20 years old had higher score on psychological health (3.67), On the other hand, in COPING strategies, participants in all the age groups had similar higher score on emotion focused COPING strategies. Fig. 2 (B) described the gender-based quality of life among the participants where female and male had higher similar score on physical health (3.43) and in COPING strategies male and female both had the similar high mean score on emotion focused COPING (F-14, M-14). Fig. 2 (C) married participants had higher mean score (Mean-3.43) on physical health, unmarried respondents had higher mean score (Mean 3.50) on psychological health. COPING strategies participants who were married and unmarried had similar high mean score (14). Fig.2 (D) Unemployed had the highest mean score on physical health (4.39); On coping strategies, students, health care professionals, law enforcement agency, housewife, government employer, private jobs, farmers and unemployed all had quite similar mean score on emotion focused coping.

**Figure 1:**
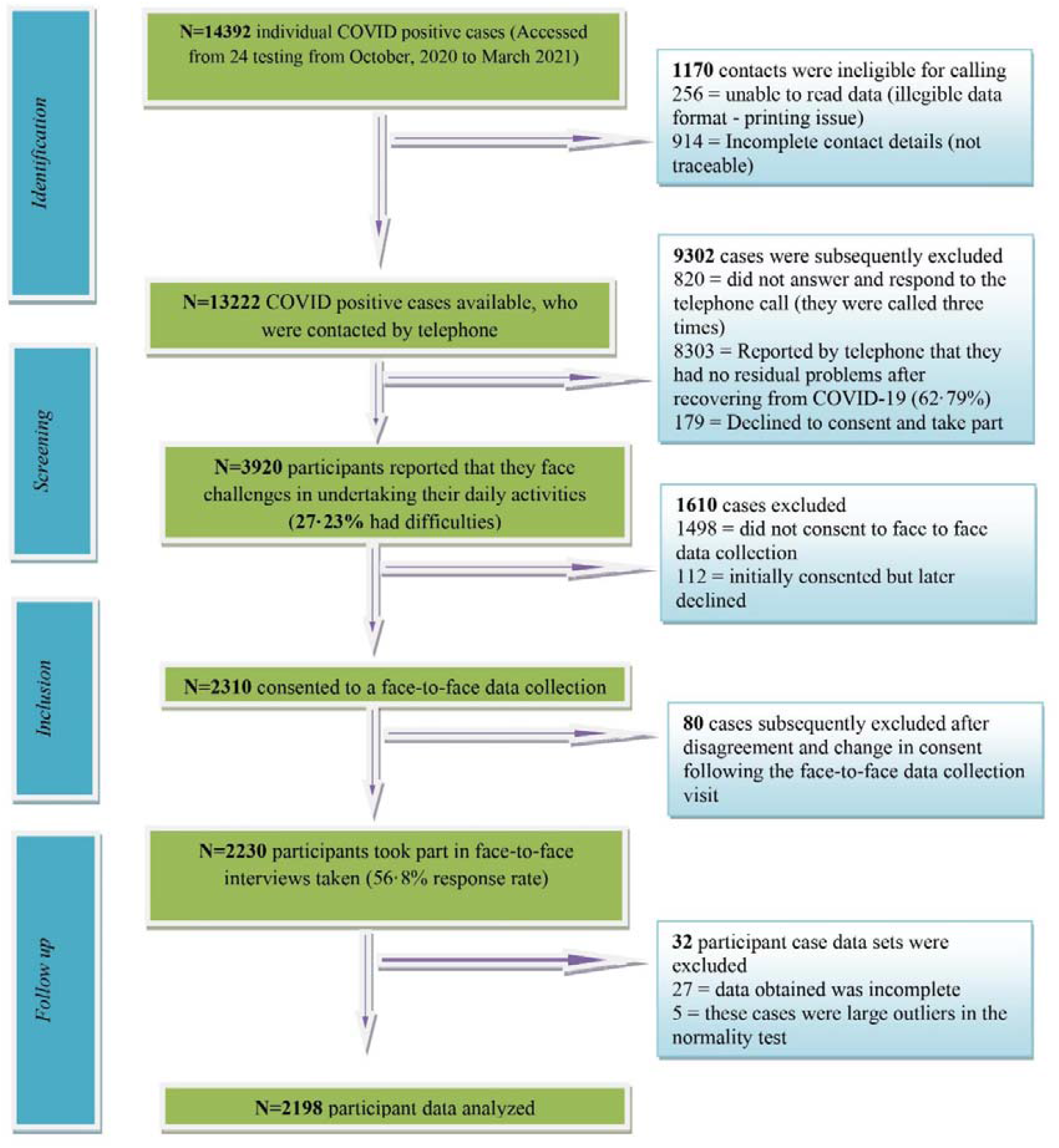
STROBE flow diagram of the study

**Figure 2:**
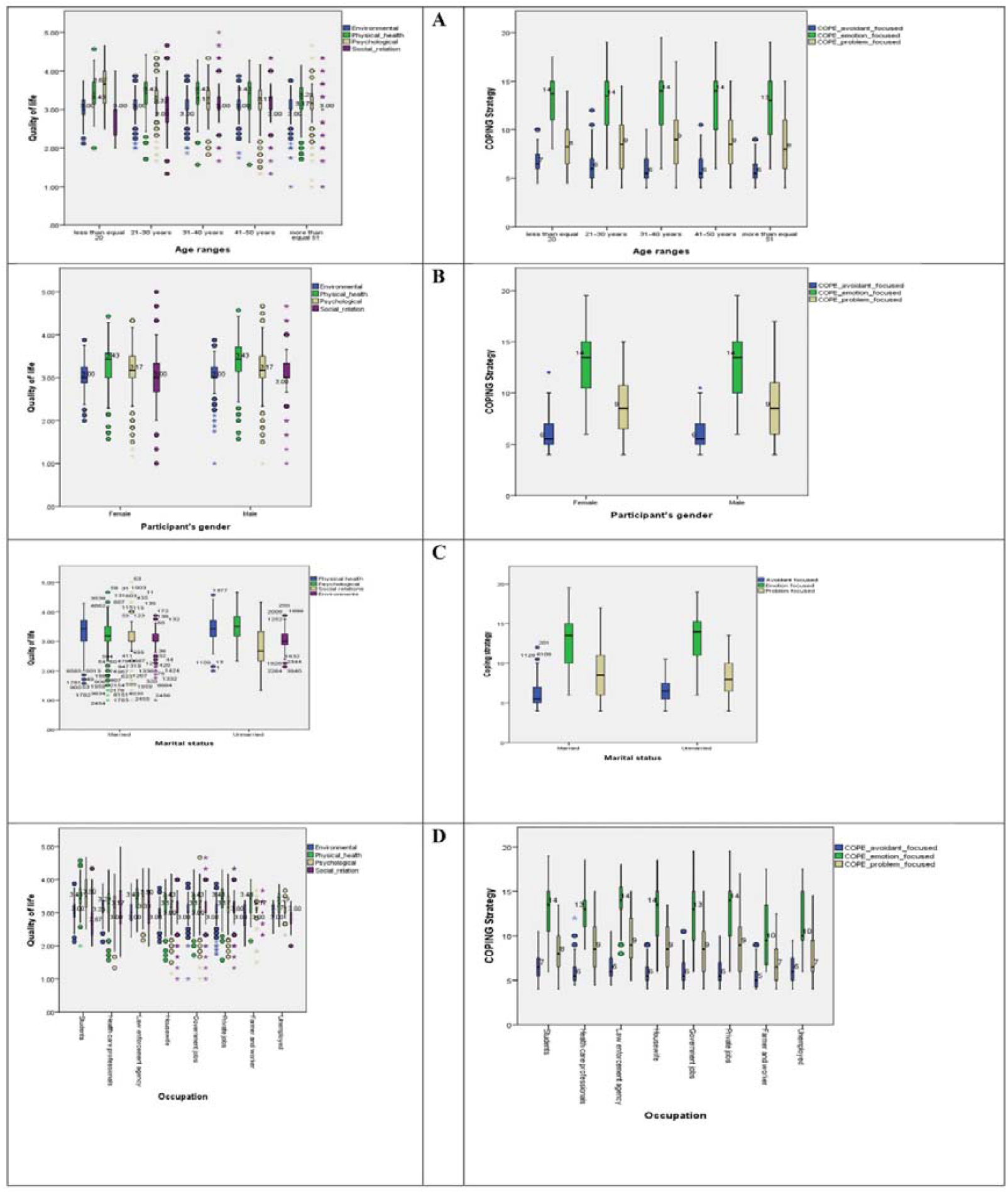
Box plot of COPING and QoL.

## Discussion

The demographic statistics showed most of the participants were in their third and fourth decade of life 32.48% (n=714), and that majority of the participants were 72.38% (n=1591). A study from China showed similar findings, where males were more affected by COVID-19 than females [24]. The prevalence (68.70%) of COVID-19 was found to be higher in the semi-urban areas, meaning their residential area was in the district or sub district of Upazila Level. Education data showed that out of 2198 participants, half of the sample 50.09% (n=1101) completed their Bachelor’s Degree at minimum, and participants involved in business as an occupation included a large proportion at 52.96% (1164). Similar demographic characteristics were evident from another article where most of the participants were in their third and fourth decade of life, males, completed Bachelor’s Degree and residing in an urban area [25]. From the total data, 48.11 percent had co-morbidities, with hypertension accounting for the highest 12.33 percent and diabetes mellitus accounting for the second highest 10.92 percent. Symptom-responses were more prevalent, in our study, among those with higher education and mainly businessmen.

Furthermore, this community-based study showed coping strategies and QoL for COVID-19 survivors’ of 2198 participants derived from both rural and urban area from the eight divisions of Bangladesh. It represents, age ≥ 20 years old, and shows a significant correlation with avoidant coping strategies which indicates their physical or cognitive efforts to disengage from the stressor. Additionally, a low mean score with the age of 41-50 years indicative of adaptive coping. Problem focused coping showed a significant correlation with male gender. However, female was more prone to emotion focused coping in this study. This possible reason is related to the accepted cultural expectation in Bangladesh that female gender identity is connected to intuitive and emotional style coping mechanisms and cumulative burden for everyday stressors [26] Previous evidence showed that during the SARS outbreak, more women than men sought counseling for emotional reasons [27]. Also, marital status had a significant and strong correlation with avoidant coping with a mean score higher for widow or widower compared to all other categories. In the current study, education, occupation, living area and administrative division found significant co-relation with problem, emotion and avoidant coping strategies. Furthermore, higher mean score on problem focused associated with higher education levels, house-wife, urban area and Chittagong division. According to previous research, higher education is associated to more positive coping methods, regardless of gender. However, women, either married, single or separated/divorced, overall, reported a higher burden of work during COVID-19, resulting in more negative coping strategies and poorer health outcomes [28]. Family member’s shows significant co-relation with problem and emotion focused coping, whereas unaffected family members reported high mean scores related to problem solving approach. But, affected family members represented regulation of emotions that were associated with specific stressful situations. Previous studies showed that family was negatively associated with coping strategies [16].

Quality of Life (QoL) is a well-known term used by health care experts all over the world to assess for any disease outcome. COVID-19 has a significant impact on people’s health-related QoL [29]. A significant correlation found between sociodemographic and QoL of COVID-19 survivors. Age showed significant correlation with physical, psychological and social relationships, but was not significantly correlated with the environment. The highest mean score showed people living in their third decade of life, had better physical health outcomes while the age group 20 and below had better psychological and social relationship status. Gender data showed men reported slightly better QoL in all four domains than women. This was similar to a previous Jordanian study where women reported higher rates of depression and lower QoL compared to men [29]. Marital status reported that unmarried participants have better psychological and social relationship than married participants. Similar findings were found in a previous study which stated that marriage initiated a process of increasing reliance on and time spent with the partner and family relatives. Furthermore, it resulted less reliance on and time with friends and non-relatives’ peers so, married people had tend to participate in fewer and more family-focused activities rather than social activities. [30] Bachelor or above degree holders showed better physical, psychological and environmental quality of life; whereas, people with no formal education represented better social relationship score. Previous studies showed higher levels of knowledge and education were all linked to a positive attitudes and health preventive practices during COVID-19 [31]. Occupation mean score showed students had significant physical, psychological, social and environmental score which means in Bangladesh student poses overall good quality of life. Though evidence from developed country shows students has negative impact with quality of life during post COVID-19 situation [32]. Psychological and social relationship scores showed significant relationship with people who living in urban areas. In terms of administrative divisions, people living in Chittagong reported higher psychological, social and environmental mean score than all other divisions. Respondents with no reported comorbidities also showed better physical, psychological and environmental scores compare with respondents with at least one or more comorbidities.

Correlation between COPING and QoL showed problem focused coping strategies were positively correlated with psychological and social relations which indicate they are able to manage more stressful situations. Emotion focused showed positive correlation with psychological and social realms, but were negatively associated with physical health and environmental health, indicating they are capable of regulating emotions associated with the stressful situation. Avoidant focused coping strategy was positively correlated with psychological health indicating better stressor coping skills; however, it was also negatively associated with physical and environment, indicative of a more adaptive coping strategy. Another study showed individuals who employed an avoidance coping technique had lower levels of wellbeing and QOL, which is often considered a maladaptive coping strategy [31]. From the box plot it shows the second decade of life had a higher mean score with physical health, and all age groups had a higher mean score with emotion focused. Both male and female QoL scores were identical in terms of physical health, but coping strategy represents better score in terms of emotional health. Previous evidence showed women poses more negative impact on the psychological health compare with men [32]. Married individuals had a higher mean score for physical health and unmarried participants had a higher mean score for psychological health, but all participants had a high mean score for avoidant coping. Another study showed individuals that were unmarried or divorced also reported a lower quality of life, possibly related to cognitive stressors [33].

## Conclusion

According to our study, during the COVID-19 pandemic fourth wave, where long periods of quarantine and lockdown were enforced across Bangladesh, there was a high report of anxiety and poor coping strategies which was directly related to psychological, emotional, physical and cognitive health outcomes and decreased quality of life for respondents across the eight Districts in Bangladesh. In addition, our study identified higher prevalence of COVID-19 in semi-urban areas. Further research could be done to find the causative reason behind the higher prevalence rate. Education, occupation and living area showed problem focused coping strategy. Education plays an important role and higher education is associated to more positive coping methods. Men had a higher quality of life and were more problem-oriented, whereas women were more emotion-focused in their coping. The government should place a greater emphasis on education and women’s health and advocate on health promotion strategies for the vulnerable female population

## Data Availability

Data will be available on request and was shared anonymously or
following ethical principle and made available to others according to the necessity of the
study. Data was available at a public repository

https://www.kaggle.com/datasets/rubayetshafin/qol-coping-covid-positive

## Declaration

### Ethics approval and consent to participate

Ethical permission was obtained from The Institutional Review Board, Institute of Physiotherapy Rehabilitation and Research, BPA (Ethical review committee at Bangladesh Physiotherapy Association) on September 19, 2020 (BPA-IPRR/IRB/16/06/2020/086) and CRP Ethics Committee, Centre for the Rehabilitation of the Paralysed (CRP) on October 10,2020 (CRP-R&E-0401-336) respectively. The study was registered retrospectively at World Health Organization (WHO) Primary Clinical trial registry platform (CTRI/2020/09/028165) on 30/09/2020 with the title “Symptoms presentation among the COVID-19 survivors in Bangladesh”. Written approval for data collection obtained from the Directorate General of Health Services (DGHS) of the Ministry of Health and Family Welfare in Bangladesh (Supplementary File A). Verbal consent was obtained during the initial telephone call and written consent was obtained at interview. The principles of the Helsinki Declaration [34] were followed throughout the research to ensure confidentiality, ethics and privacy.

### Consent for publication

Not Applicable

### Availability of data and materials

Data will be available on request and was shared anonymously or following ethical principle and made available to others according to the necessity of the study. Data was available at a public repository (https://www.kaggle.com/rubayetshafin/qol-coping-covid-positive).

### Competing interest

The authors declared that they have no competing interests regarding publication of this journal.

### Funding

This was self-funded research. Authors did not receive any kind of fund to conduct this research and all the abroad scholar’s participation in this research voluntarily.

### Authors Contribution

MAH, RS, MSA, MSR, IKJ conceptualised the study. MAH, LMW, VR, RS, MSR, MSA, IKJ curated and collated the data. MAH, LMW, VR, RS, MSH, MFK, MSR, MSA, MDH, FSR, IKJ did validation. RS, MSA, TA, MRI visualised the data. MAH, LMW, VR, RS, MSH, MFK, AHR, MSR contributed to the model development. RS, MSA, TA, MRI did the formal analysis. RS, MSA, MSR, MAH wrote the manuscript. LMW, VR, MSA, MSR, IKJ, MSH. MFK, MAH, MNH, MDH, FSR reviewed the manuscript. All author had full access to all the data in the study and the corresponding author had final responsibility for the decision to submit for publication.

## Acknowledgement

Authors acknowledges the assessor for their tremendous effort to collect the data from all over the Bangladesh.

## Notes

### Competing Interest Statement

The authors have declared no competing interest.

### Funding Statement

This study did not receive any funding

### Author Declarations

Ethical permission for this study was obtained from the CRP Ethics Committee, Centre for the Rehabilitation of the Paralysed (CRP) on October 10,2020 (CRP-R&E-0401-336)

